# Importance of adequate COVID-19 case definitions in the SARS-CoV-2 pandemic

**DOI:** 10.1101/2021.06.13.21258845

**Authors:** Isaac Núñez, Yanink Caro-Vega, Pablo F. Belaunzarán-Zamudio

**Author notes:** **Corresponding autor:** Isaac Núñez. Instituto Nacional de Ciencias Médicas y Nutrición Salvador Zubirán, Vasco de Quiroga #15, Tlalpan Mexico City, Mexico, postal code 14080.

## Abstract

**Background:** Epidemiologic case definitions serve a myriad of purposes during a pandemic, including contact tracing and monitoring disease trends. It is unknown how any COVID-19 case definition fares against the current gold standard of molecular or antigen tests.

**Methods:** We calculated the diagnostic properties of five COVID-19 definitions (three of the Mexican government and two of the WHO) using open data of suspected COVID-19 cases in Mexico City from March 24th 2020 until January 31st 2021.

**Results:** All 1,632,420 people included in the analysis met the WHO suspected case definition (sensitivity 100%, specificity 0%). The WHO probable case definition was met by 1.4%, while the first and second Mexican suspected case had sensitivities of 61 and 62% and specificities of 58 and 62%, respectively. Confirmed case by epidemiological contact had a low sensitivity (33%) but slightly higher specificity (77%).

**Conclusions:** Case definitions should maximize sensitivity, especially in a high-transmission area such as Mexico City. The WHO suspected case definition has the potential for detecting most symptomatic cases. We underline the need for routine evaluation of case definitions as new evidence arises to maximize their usefulness.

## Introduction

Epidemiologic case definitions for a disease are vital in surveillance during epidemics. In this context, the aim of a case definition is to be highly sensitive as to miss the fewest true cases of the disease. In low-income countries, case definitions are invaluable to make decisions on isolation, contact tracing, and monitoring disease trends, since definitive tests might be scarce, unavailable, and highly expensive [1]. During the COVID-19 pandemic the World Health Organization (WHO) released its recommended version of case definitions for suspected, probable and definitive COVID-19 cases, which have been periodically updated [2]. Countries have also released their own definitions with irregular updating [3, 4], even though case definitions must be updated according to new scientific evidence as to increase their diagnostic value [1-4].

Mexico released the first version of its COVID-19 suspected case definition in March 2020 with the aim of determining who should be tested. Only one in ten ambulatory suspected COVID-19 patients would be tested, as well as all hospitalized ones. An update in the case definition was published in august 2020, with only minor changes but the testing strategy remained the same [3-4]. Considering this deliberate under testing, suspected case definitions become especially important to account for disease undercounting, initiate contact tracing (which has not been a feature of Mexico’s pandemic response, but is elsewhere), and starting individual treatment. An important caveat is that the people who design these definitions are in many cases not the same people that have to apply them on the field. Thus, a dissociation could occur between intended and actual use.

Mexico City is a large metropolitan area with 9,209,944 habitants [5]. In this analysis we calculated the diagnostic properties of the definitions of COVID-19 cases of the Mexican Ministry of Health (MoH) and those of the WHO, to determine their adequacy for epidemiological monitoring purposes.

## Materials and Methods

We used open data from the Mexico City government for reported cases of suspected COVID-19 between March 24th 2020 and January 31st 2021 [6]. We calculated the diagnostic properties (sensitivity, specificity, positive predictive value, negative predictive value, positive likelihood ratio, and negative likelihood ratio), as well as post-test probabilities of five different epidemiological COVID-19 case definitions: three issued by the MoH for COVID-19 surveillance purposes (suspected case, updated suspected case definition, and suspected case “confirmed” by epidemiological linkage to a laboratory confirmed case) and two WHO recommended definitions (suspected and probable) [2-4]. A comparison between these definitions is provided in Table A.1. The COVID-19 suspected case definition of the MoH was issued in March 2020 and included anybody seeking care for at least one of the following symptoms starting within the 7 previous days: cough, dyspnea, fever or headache; with at least one of the following: myalgia, arthralgia, sore throat, thoracic pain, rhinorrhea, polypnea or conjunctivitis [3]. COVID-19 suspected case definition was updated by the MoH in August 2020 (adding chills, anosmia and dysgeusia and expanding the period of symptoms onset from 7 to 10 days) [4]. The case definition for COVID-19 confirmed by epidemiological linkage to a laboratory confirmed case (anyone meeting the COVID-19 suspected case criteria that have had contact with a laboratory confirmed case within the previous 14 days) [4]. We substituted contact with “confirmed case” with contact with an “individual with respiratory symptoms”, as only information on this variable was available [6]. The MoH COVID-19 case definition is the same case definition used for surveillance activities of seasonal Influenza thus, tests are only performed to symptomatic patients but only one in ten ambulatory patients are tested, while all hospitalized are [3]. There are no pre-established criteria or even guidelines on which ambulatory patients test for SARS-CoV-2, and decisions about testing depends heavily in clinical judgement and tests availability on sentinel sites. The revised WHO COVID-19 case definitions for suspected (which includes a set of different options of clinical and epidemiological criteria, as shown in Table A.1) and probable cases (which requires the presence of the clinical criteria in suspected cases (acute onset of fever and cough OR acute onset of any three or more of the following symptoms: fever, cough, general weakness/fatigue, headache, myalgia, sore throat, coryza, dyspnea, anorexia/nausea/vomiting, diarrhea, altered mental status in combination) in combination with chest imaging showing findings suggestive of COVID-19 disease, which we replaced with the variable “clinical diagnosis of pneumonia”, since no chest imaging variable was available in the database [2-4].

Since all tested patients in Mexico are symptomatic, the diagnostic gold standard for our calculation of the diagnostic properties of case definitions was having either a positive molecular test (real-time reverse-transcriptase polymerase chain reaction; RT-PCR) or a positive antigen test. Post-test probabilities of COVID-19 were calculated using the daily proportion of positive molecular or antigen tests and graphed using 7-day rolling means. Data is freely available at the official Mexico City government COVID-19 website and the R code used for the analysis will be made freely available with the final version of the article [6].

## Results

A total of 1 725 514 people were registered in the Mexico City open database during the study period. We excluded 64 967 (3.8%) that had no test result and 28 127 (1.6%) that had lost or unprocessed RT-PCR tests with no available antigen test. Thus, we included 1 632 420 patients in the analysis. There were 502 041 (30.7%) positive, and 1 129 923 (69.2%) negative RT-PCR for SARS-CoV-2, and 456 (0.03%) had a positive test result for another virus. Antigen tests were done in 786 046 (48.1%) patients, with 204 874 (26%) positive and 641 500 (74%) negative results.

Both Mexican MoH definitions of suspected COVID-19 cases had similar diagnostic properties, with slightly better characteristics in the updated definition (Table 1). All patients met the WHO definition of suspected COVID-19 case, with a perfect sensitivity, specificity of 0%, positive predictive value of 31%, and negative predictive value of 0%. Meanwhile, the WHO case definition for probable COVID-19 was met by very few patients (23 417, 1.4%), showing a sensitivity of 3%, specificity of 99%, PPV of 51%, and NPV of 71%. It is noteworthy that all patients that did not meet the probable case definition met the suspected case definition.

**Table 1.** Diagnostic properties of COVID-19 epidemiological case definitions in Mexico City

| Definition |  | Positive RT-PCR or antigen test | Negative RT-PCR or antigen test | Properties |  |
| --- | --- | --- | --- | --- | --- |
| Suspected case Mexican MoH (first) | Yes | 306 727 | 474 933 | Sens | 61% |
|  |  |  |  | Spec | 58% |
|  |  |  |  | PPV | 39% |
|  | No | 195 314 | 655 446 | NPV | 77% |
|  |  |  |  | LR+ | 1.45 |
|  |  |  |  | LR- | 0.67 |
| Suspected case Mexican MoH (updated) | Yes | 310 326 | 423 921 | Sens | 62% |
|  |  |  |  | Spec | 62% |
|  |  |  |  | PPV | 42% |
|  | No | 191 715 | 706 458 | NPV | 79% |
|  |  |  |  | LR+ | 1.63 |
|  |  |  |  | LR- | 0.61 |
| Confirmed case Mexican MoH (epidemiological contact) | Yes | 164 232 | 256 328 | Sens | 33% |
|  |  |  |  | Spec | 77% |
|  |  |  |  | PPV | 39% |
|  | No | 337 809 | 874 051 | NPV | 72% |
|  |  |  |  | LR+ | 1.43 |
|  |  |  |  | LR- | 0.87 |
| WHO suspected case | Yes | 502 041 | 1 130 379 | Sens | 100% |
|  |  |  |  | Spec | 0% |
|  |  |  |  | PPV | 31% |
|  | No | 0 | 0 | NPV | 0% |
|  |  |  |  | LR+ | 1 |
|  |  |  |  | LR- | * |
| WHO probable case | Yes | 15 351 | 8 066 | Sens | 3% |
|  |  |  |  | Spe | 99% |
|  |  |  |  | PPV | 66% |
|  | No | 486 690 | 1 122 313 | NPV | 70% |
|  |  |  |  | LR+ | 3 |
|  |  |  |  | LR- | 0.98 |

LR+: positive likelihood ratio; LR-: negative likelihood ratio; Mx: Mexico; NPV: negative predictive value; PPV: positive predictive value; RT-PCR: real time polymerase chain reaction; Sens: sensibility; Spec: specificity; WHO: World Health Organization

Post-test probability varied greatly according to the pre-test probability and the definition utilized, with a mean probability of 44% (SD 10) for the first MoH case definition, 46% (SD 10) for the updated definition, 43% (SD 10) for the MoH definition of confirmed case by epidemiological contact, 35% (SD 9) for the WHO definition of suspected case, and 61% (SD 10) for the WHO definition of probable case. Post-test probability along the study period for each definition is shown in Figure 1.

**Figure 1.**
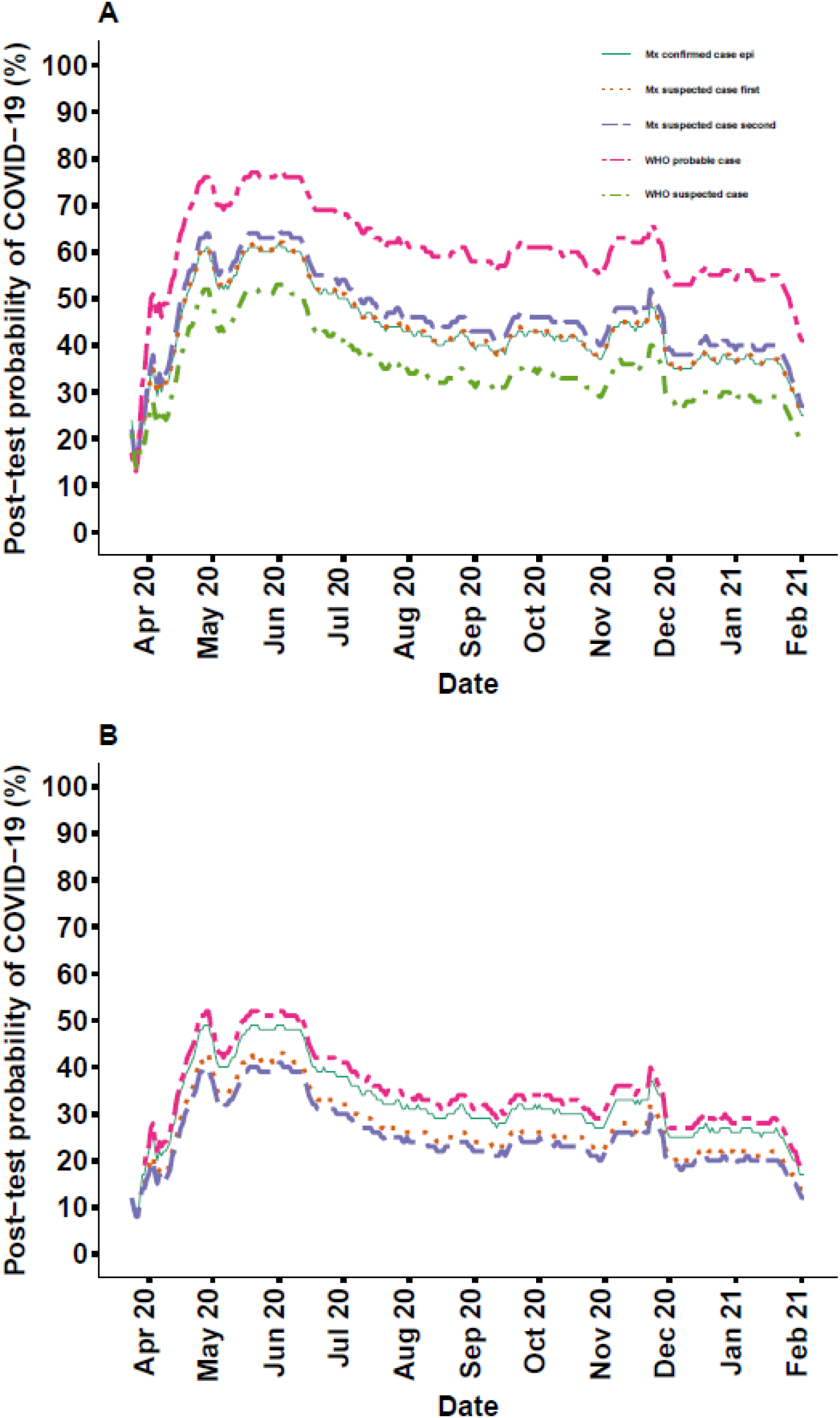
Post-test probability of COVID-19 according to several case definitions. A) shows post-test probabilities in case of meeting a given case definition. B) shows post-test probabilities in case of not meeting the case definition. Mx confirmed case epi: Mexico definition for confirmed case by epidemiological contact; Mx suspected case first: Mexico first definition for suspected case; Mx suspected case first: Mexico second definition for suspected case; WHO probable case: WHO definition for probable case; WHO suspected case: WHO definition for suspected case. Gold standard was considered to be either a positive molecular or antigen SARS-CoV-2 test. As the WHO suspected case definition had a negative predictive value of 0%, it does not appear in B).

## Discussion

Epidemiological case definitions are indispensable for surveillance but are riddled with challenges. When tallying disease cases according to case definition, changing it can increase the number of cases can increase several times, as described by Tsang et al. with COVID-19 [7]. We observed that the three COVID-19 case definitions used by the MoH have poor sensitivity (33 to 62%) in contrast with the WHO suspected case definition. This has the obvious implication that the suspected case definition of the MoH is not being used as intended (as a screening test to decide who should be considered for testing).

Considering that theoretically it should have a sensitivity of 100%, it is fortunate that it is not being used as planned, as almost 40% of currently observed cases would be missed. A suspected case definition that is not met by many confirmed cases is not useful, for epidemiologic purposes or otherwise. Our analysis underlines the importance of this, as Mexico is a country that tests a small percentage of symptomatic people. In our context, suspected cases based on symptoms should include all but asymptomatic individuals as the WHO suspected case definition does, and be formally counted and included in epidemiologic surveillance, as most do not have access to confirmatory tests.

Thus, the WHO suspected case definition appears to be the most useful in our study population (which has heavy community transmission), and guiding testing decisions in Mexico City using it would reduce sub-estimation case count. We consider it reasonable that results would be similar if we replicated the analysis country wide, unfortunately we do not have the data to do so.

As only symptomatic people are being tested, clinical judgement remains key and patients should be retested in case of a negative result if prevalence remains high [8]. Point-of-care antigen tests might be very useful in these contexts, as their low cost allows for repeated testing [8].

Our results differ to those of a previous study with a smaller sample size where a higher sensitivity for the original MoH suspected case definition was observed (87%) [9]. Their study population was highly selected, as it included patients from a single healthcare system from a single city that had exhaustive clinical information, including a complete medical history, which could explain the higher sensitivity.

Our study has several limitations. We used repurposed data that did not have information on several variables, such as anosmia, dysgeusia, and radiological imaging. The incidence of anosmia and/or dysgeusia in Mexican COVID-19 patients is unknown, but elsewhere it has been reported of 35% [10]. This could improve the sensitivity of the second MoH definition. Only one in ten ambulatory patients are tested, and these patients could differ in important ways that we are unable to account for, such as subjective disease severity.

Furthermore, false negative tests are well known and limit our definition of gold standard [11-13]. This is especially important given the high post-test probability observed throughout the study period (>10%). Accounting for false negative tests would increase the post-test probability, and thus a negative test would not rule out the disease in high prevalence areas such as this. Our analysis supports the fact that case definitions should be formally evaluated as to ensure their usefulness.

## Data Availability

All data utilized is openly available in the cited government webpages. Code used will be made freely available with the final version of the article.

## Acknowledgments

We would like to thank the government of Mexico City for their open data policy. Also, we give our gratitude to healthcare workers. Your work makes the difference.

## Funding

We received no funding for this study.

### Abbreviations and acronyms

COVID-19: Coronavirus disease 2019
WHO: World Health Organization
MoH: Mexican Ministry of Health
Mx: Mexico

