## Supplementary material for "Importance of adequate COVID-19 case definitions in the SARS-CoV-2 pandemic": Table A.1

Appendix Table 1. Comparison of Mexican Ministry of Health (MoH) and World Health Organization (WHO) COVID-19 Case definitions

| **Mexican Ministry of Health definitions** | | | **WHO definitions** | |
| --- | --- | --- | --- | --- |
| **Suspected case (March 24, 2020 definition)** | **Suspected case (August 25, 2020 definition)** | **Confirmed case by epidemiological link (August 25, 2020 definition)** | **Suspected case of SARS-CoV-2 infection** | **Probable case of SARS-CoV-2 infection** |
| Any person that presented in the last seven days any one of these symptoms: cough, dyspnea, fever or headache **AND** at least one of the following:  -Myalgias  -Arthralgias  -Sore throat  -Chest pain  -Rinorrhea  -Polypnea  -Conjunctivitis | Any person that presented in the last ten days any one of these symptoms: cough, dyspnea, fever or headache **AND** at least one of the following:  -Myalgias  -Arthralgias  -Sore throat  -Chills  -Chest pain  -Rinorrhea  -Polypnea  -Conjunctivitis  -Anosmia  -Dysgeusia | Any person that presented in the last ten days any one of these symptoms: cough, dyspnea, fever or headache **AND** at least one of the following:  -Myalgias  -Arthralgias  -Sore throat  -Chills  -Chest pain  -Rinorrhea  -Polypnea  -Conjunctivitis  -Anosmia  -Dysgeusia  **AND**    Contact with a laboratory confirmed COVID-19 case during the last 14 days. | Three options, A through C: A. A person who meets the clinical **AND** epidemiological criteria: Clinical criteria: 1. Acute onset of fever **AND** cough; **OR** 2. Acute onset of ANY THREE OR MORE of the following signs or symptoms: fever, cough, general weakness, fatigue, headache, myalgia, sore throat, coryza, dyspnoea, anorexia/nausea/vomiting, diarrhoea, altered mental status. **AND** Epidemiological criteria: 1. Residing or working in a setting with high risk of transmission of the virus: for example, closed residential settings and humanitarian settings, such as camp and camp-like settings for displaced persons, any time within the 14 days before symptom onset; **OR** 2**.** Residing in or travel to an area with community transmission anytime within the 14 days before symptom onset; **OR** **3.** Working in health setting, including within health facilities and within households, anytime within the 14 days before symptom onset. **B**. A patient with severe acute respiratory illness (SARI: acute respiratory infection with history of fever or measured fever of ≥ 38 C°; **AND** cough; with onset within the last 10 days; **AND** who requires hospitalization). **C.** An asymptomatic person not meeting epidemiologic criteria with a positive SARS-CoV-2 antigen-detecting rapid diagnostic test (Ag-RDT) | Four options, A through D:  **A.** A patient who meets clinical criteria of suspected case AND is a contact of a probable or confirmed case or is linked to a COVID-19 cluster.  **B.** A suspected case (described above) with chest imaging showing findings suggestive of COVID-19 disease.  C. A person with recent onset of anosmia (loss of smell) or ageusia (loss of taste) in the absence of any other identified cause.  D. Death, not otherwise explained, in an adult with respiratory distress preceding death AND who was a contact of a probable or confirmed case or linked to a COVID-19 cluster. |
